# Photodynamic therapy is a safe and feasible adjunct to percutaneous drainage of deep tissue abscesses: Results of a first in humans Phase 1 clinical trial

**DOI:** 10.1101/2023.10.16.23297086

**Authors:** Timothy M. Baran, David A. Bass, Laurie Christensen, Erica Longbine, Maria D. Favella, Thomas H. Foster, Ashwani K. Sharma

## Abstract

**Background:** Standard of care for abscess management includes image-guided percutaneous drainage and antibiotics. However, cure rates vary between patients and there is growing concern for antibiotic-resistant bacteria. Photodynamic therapy (PDT), which utilizes light-activated dyes to generate cytotoxic reactive species, could complement the standard of care by sterilizing the abscess at time of drainage.

**Purpose:** The goal of this study was to perform a first in humans Phase 1 clinical study evaluating safety and feasibility of PDT with methylene blue (MB) at the time of percutaneous abscess drainage. This was accomplished through an open-label dose escalation study, with duration of light delivery escalated from 5-30 minutes.

**Materials and Methods:** We performed MB-PDT in 18 subjects undergoing percutaneous abscess drainage. Following standard of care drainage, 1 mg/mL MB was delivered for 10 minutes. MB was aspirated, and 1% lipid emulsion infused to homogenize light dose at the cavity wall. An optical fiber was advanced to the approximate center of the abscess for 665 nm laser illumination at 20 mW/cm^2^.

**Results:** MB-PDT at the time of abscess drainage was safe and feasible in all cases, with no evidence of fat embolism due to lipid emulsion or adverse reaction to MB observed. No study-related adverse or serious adverse events were encountered, and the procedure was well tolerated by all subjects. While the study was not designed or powered to determine efficacy, time to resolution of clinical symptoms was significantly decreased in subjects receiving higher fluences (p=0.028). Additionally, drainage catheter output post-procedure was decreased in subjects receiving higher fluences (ρ=-0.18), although this difference was not significant (p=0.43).

**Conclusion:** MB-PDT is a safe and feasible adjunct to image-guided percutaneous abscess drainage. Clinical measures indicate a dose-dependent response to PDT, motivating future Phase 2 studies evaluating the efficacy of MB-PDT in this patient population.

## Introduction

Abscesses form through interaction between acute microbial infection and the host immune system, resulting in fever, nausea, and acute abdominal pain. If untreated, mortality ranges from 35-100%^1,2^. Abscesses are routinely managed by image-guided percutaneous drainage and systemic antibiotics, which has reduced mortality and morbidity relative to open surgical drainage^3,4^. Despite this, abscesses remain a major source of morbidity, mortality, and hospital stay^4^. Further, response can vary widely between patients. Particularly for abscesses that are complex or loculated, cure rates can be as low as 30%^4^.

This can result in abscess recurrence, requiring repeated drainage procedures and catheter replacement, which increases hospital stay, costs, and patient discomfort, as well as prolonged antibiotic administration. This is of particular concern, as antibiotic-resistant bacteria are present in abscesses^5–7^ and incidence of these is expected to increase^8^. The development of alternative treatment options is therefore paramount for effective management of future abscess patients.

Photodynamic therapy (PDT) is a promising treatment for cancer and infectious disease that relies upon excitation of photosensitive drugs known as photosensitizers by visible light to generate cytotoxic reactive oxygen species^9^. PDT is effective against antibiotic-resistant bacteria^10,11^, and does not result in acquired resistance^9^. With the photosensitizer methylene blue (MB), PDT has been used to treat endodontic infection^12,13^, periodontal pockets^14^, infected wounds^15^, and onychomycosis and infected diabetic foot ulcers^16^. MB-PDT was efficacious against bacteria cultured from human abscess aspirates^6^ and bacterial strains typically found in human abscesses^17^. Simulation studies showed that a large proportion of abscess patients would be eligible for this procedure^18^, with as many as 92% of patients expected to experience a therapeutic benefit^19^. However, PDT has never been used to treat infections deep in the body, such as abdominal abscesses.

Based on these encouraging pre-clinical results and the large potential clinical benefit, we initiated a prospective, open-label Phase 1 clinical trial investigating MB-PDT at the time of percutaneous abscess drainage. The goal of this study was to evaluate safety and feasibility of this procedure, as MB-PDT has not been used previously in human abscesses. We performed a dose escalation study, with MB concentration fixed and laser illumination escalated from 5-30 minutes. We hypothesized that MB-PDT would be safe and well tolerated at all illumination durations, with some evidence for efficacy apparent for longer illumination. We previously published a case report from one of these subjects^20^. Here, we report the full results of the completed Phase 1 clinical trial.

## Materials and Methods

### Regulatory Approval

All study procedures were approved by the Institutional Review Board, and subjects provided written informed consent. Data collection and handling was performed in compliance with HIPAA. This study is registered on ClinicalTrials.gov (NCT02240498).

### Study Design

The goal of this prospective, open-label Phase 1 clinical trial was to determine safety and feasibility of MB-PDT at the time of image-guided percutaneous abscess drainage. We employed a dose escalation scheme, with MB concentration and fluence rate held constant, and fluence escalated using a 3+3 study design^21^. Illumination time was escalated from 5-30 minutes in five minute increments, with three subjects treated at each.

### Participants

Participants were recruited from patients scheduled for image-guided percutaneous abscess drainage. Inclusion criteria were: (1) aged 18+ years and (2) abscess diameter <8 cm. Exclusion criteria were: (1) pregnancy, (2) lactation, (3) allergy to contrast media, narcotics, sedatives, atropine, or eggs, (4) necrotic tissue requiring surgical debridement, (5) severely compromised cardiopulmonary function, (6) thrombocytopenia, (7) uncorrectable coagulopathy, (8) poor kidney function (serum creatinine > 3 mg/dL), (9) lack of safe pathway to abscess, (10) patient unable/unwilling to undergo study procedure, (11) multiple separate abscesses, and (12) patient taking certain serotonergic psychiatric medications.

Abscesses with diameter >8 cm were excluded due to limitations on maximum laser optical power. Patients with multiple abscesses were excluded to isolate effects of MB-PDT for adverse event monitoring. Patients taking specific serotonergic psychiatric medications were excluded due to reports of MB interactions^22^.

### Study Procedures

All subjects first received standard of care image-guided percutaneous drainage under CT or ultrasound guidance. Syringe aspiration of abscess material was collected and sent to the Clinical Microbiology Laboratory for identification and antibiotic susceptibility testing.

Using the same access used for drainage, sterile 1 mg/mL MB (BPI Labs, LLC, Largo, FL) was infused into the abscess. The volume infused was matched to the volume of material aspirated during drainage. Following 10 minute incubation, during which MB was taken up by remaining bacteria within the abscess, MB was aspirated and the cavity flushed with sterile saline. The abscess was filled with sterile 1% lipid emulsion (Intralipid (Baxter Healthcare, Deerfield, IL) or Nutrilipid (B. Braun Medical Inc., Bethlehem, PA)) to gently distend the cavity and homogenize light dose through scattering.

A sterile optical fiber (Vari-Lase Platinum Bright Tip, Vascular Solutions, Minneapolis, MN) connected to a 665 nm diode laser system (ML7710, Modulight, Tampere, Finland) was advanced to the approximate abscess center. Laser power was set to achieve a 10-20 mW/cm^2^ fluence rate, based upon abscess dimensions on pre-procedure CT imaging. Light was delivered for 5-30 minutes, resulting in fluences of 6-36 J/cm^2^. The optical fiber was then withdrawn, lipid emulsion aspirated, and the cavity flushed with sterile saline. Administered and aspirated volumes of MB and lipid were recorded.

Optical spectroscopy measurements were additionally performed before and after MB administration. Preliminary results were reported previously^23^, with full results described in a separate publication^24^.

### Outcome Evaluation

The primary outcome was safety, evaluated by absence of: (1) fat embolism, (2) MB escape with evidence of adverse reaction, (3) disruption of abscess wall or damage to surrounding tissue, and (4) need for surgery to remove broken optical fiber.

Fat embolism has been reported as a side effect of lipid emulsion administered intravenously at high concentration. Side effects related to MB have been reported at intravenous doses of 4 mg/kg or greater. Subjects with a difference between administered and aspirated MB volumes which could result in a dose >3 mg/kg were monitored with CO-oximetry for methemoglobin formation.

While the study was not designed or powered to evaluate efficacy, preliminary evaluation was inferred through clinical endpoints. These included: (1) time to drainage catheter removal, (2) drainage catheter output, and (3) time to clinical symptom resolution. Resolution of clinical symptoms was defined as the first day post-intervention where the subject was afebrile, reported no pain/malaise, and was able to have bowel movement.

Subjects were followed daily until drainage catheter removal, for a minimum of 48 hours. Drainage catheters were removed when output reduced to 5 cc/day. Secondary endpoints were evaluated by review of subjects’ electronic health record. All subject data were reviewed by an Independent Safety Monitor.

### Statistical Analysis

Continuous values are summarized as mean±standard deviation, categorical values are summarized as proportion (95% confidence interval). Outcomes were compared using unpaired t-tests or ordinary one-way ANOVA, with Tukey’s test for multiple comparisons. Correlations were assessed with Spearman correlation coefficients. Analyses controlling for abscess volume utilized linear regression. P values <0.05 were considered significant. All analyses were performed by one of the authors (TMB) using SPSS (v28.0, IBM Corp, Armonk, NY) and GraphPad Prism (v6.07, GraphPad Software, Inc., Boston, MA).

As this was a Phase 1 safety/feasibility study, effect sizes for hypothesized efficacy were not used to determine sample size. Instead, a 3+3 study design was utilized as described above.

## Results

### Participant Characteristics

18 subjects were treated, with demographics summarized in Table 1. Race and ethnicity information were extracted from subjects’ electronic health record. A total of 56 subjects were screened (see Figure 1). The most common exclusion criteria were abscess diameter >8 cm (n=11), taking serotonergic psychiatric medications (n=10), and presence of multiple abscesses (n=9).

**Figure 1:**
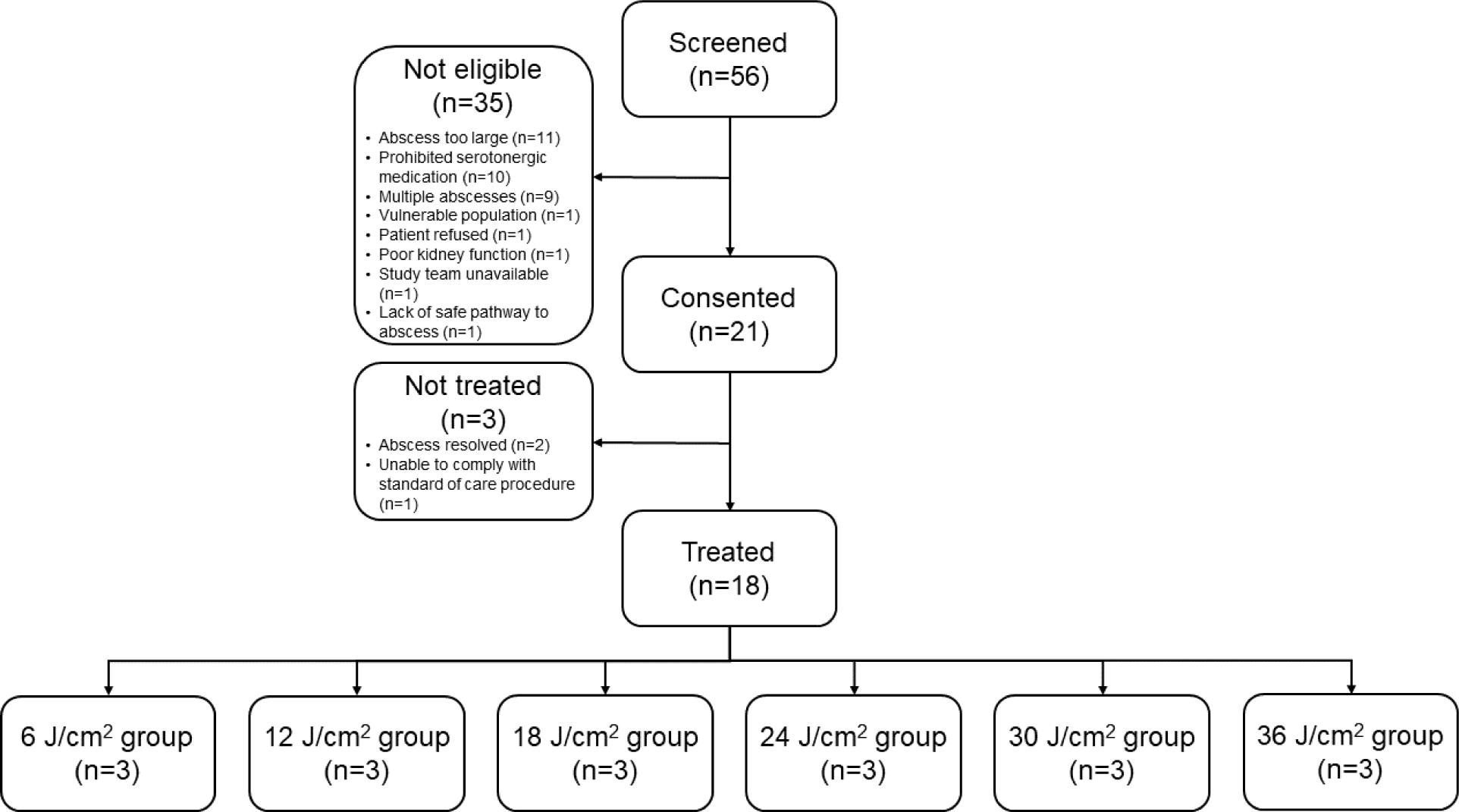
Flowchart illustrating recruitment of treated cohort.

**Table 1.** Subject demographics. * Mean ± standard deviation (range) ^†^ Proportion (95% confidence interval)

|  | Category | Summary |
| --- | --- | --- |
| Age (years)* | - | 60.1±18.3 (18-86) |
| Sex<sup>†</sup> | Female | 55.6% (30.8-78.5%) |
|  | Male | 44.4% (21.5-69.2%) |
| BMI (kg/m<sup>2</sup>)* | - | 29.3±5.9 (19.6-39.1) |
| Race<sup>†</sup> | White | 88.8% (65.3-98.6%) |
|  | Black | 11.1% (1.4-34.7%) |

### Safety and Feasibility Endpoints

No study-related adverse or serious adverse events were encountered, and MB-PDT was well-tolerated by all subjects. One subject developed gastric bleeding after intervention. However, this was determined to be unrelated to the study by both the Independent Safety Monitor and study team. This bleeding was due to an unrelated perforated duodenal ulcer at a site remote to the treated abscess.

There was no evidence of fat embolism due to lipid emulsion or adverse reaction to MB observed in any subject. 98.7% ± 3.1% (90-100% range) of infused MB was aspirated, and 96.9% ± 5.0% (83-100% range) of infused lipid emulsion was aspirated. Even if all un-aspirated MB were released into systemic circulation, the maximum whole body MB dose was 0.1 mg/kg. This is well below the 4 mg/kg threshold for potential side effects, so CO oximetry post-PDT was not required in any cases.

Criteria for technical success included successful administration of MB and lipid emulsion, placement of the optical fiber, laser irradiation, and removal of the entire optical fiber. These criteria were met in all cases, indicating that MB-PDT is technically feasible. Depending on treatment group, addition of PDT to drainage increased procedure time by ∼20-45 minutes (10 minutes MB incubation + 5-30 minutes laser illumination + ∼5 minutes for laser calibration and MB/lipid dilution).

The treatment fiber was removed intact in all cases, and post-procedure imaging showed no evidence of abscess wall disruption (see Figure 2). As no study-related adverse events were encountered, no treatment groups were repeated and no dose de-escalation was required under the 3+3 design.

**Figure 2:**
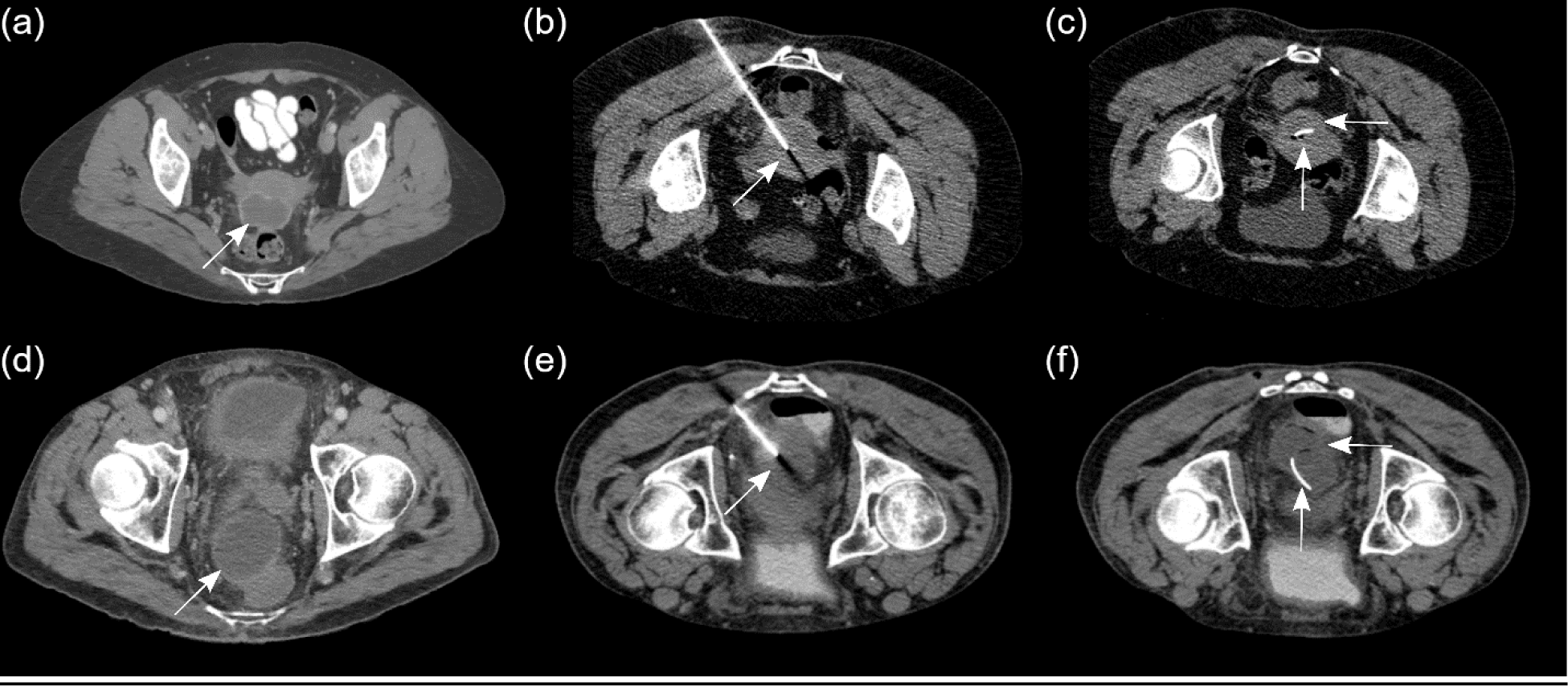
Representative (a) pre-procedure, (b) intra-procedure, and (c) end of procedure images for a subject receiving 10 minutes of laser illumination, as well as representative (d) pre-procedure, (e) intra-procedure, and (f) end of procedure images for a subject receiving 30 minutes of laser illumination. Arrows in (a) and (d) indicate the position of each subject’s pelvic abscess. Arrows in (b) and (e) indicate the distal tip of the needle used for access. Vertical arrows in (c) and (f) indicate the position of the placed pigtail catheter, while horizontal arrows indicate the intact abscess wall.

### Representative Cases

Two representative cases are shown in Figure 2. The subject shown in Figures 2a-c received 10 minute illumination (12 J/cm^2^), while the subject shown in Figures 2d-f received 30 minute illumination (36 J/cm^2^). In both cases, a well-defined pelvic abscess was visualized on pre-procedure CT imaging (Figures 2a and 2d), with image-guided percutaneous drainage performed under CT guidance (Figures 2b and 2e). Imaging after completion of PDT at the conclusion of the procedure (Figures 2c and 2f) showed no evidence of abscess wall disruption, with *in situ* drainage catheter placement shown. The subject shown in Figures 2d-f displayed more rapid abscess resolution, potentially due to administration of a larger fluence.

### Abscess Response

Of the 18 abscesses treated, only one required an additional procedure to exchange the initially placed drain. This abscess was the first treated, and received a fluence of 6 J/cm^2^ (5 minute illumination). This fluence is below the level where we have shown antimicrobial efficacy *in vitro*^6,17^, and was meant to be a conservative starting point for dose escalation. It is therefore not unexpected that response to PDT would be less pronounced in this illumination group.

Two abscesses, one in the 12 J/cm^2^ group and one in the 24 J/cm^2^ group, were not completely resolved on subsequent imaging at 4 months and 1 month post-PDT. However, these both resolved without any additional procedures, potentially indicating delayed response to intervention.

### Preliminary Efficacy Assessment

Although the study was not designed or powered to evaluate efficacy amongst treatment groups, we tracked surrogate measures of PDT response. These included drainage catheter output post-intervention, time to catheter removal, and resolution of clinical symptoms. These outcomes are summarized in Figure 3.

**Figure 3:**
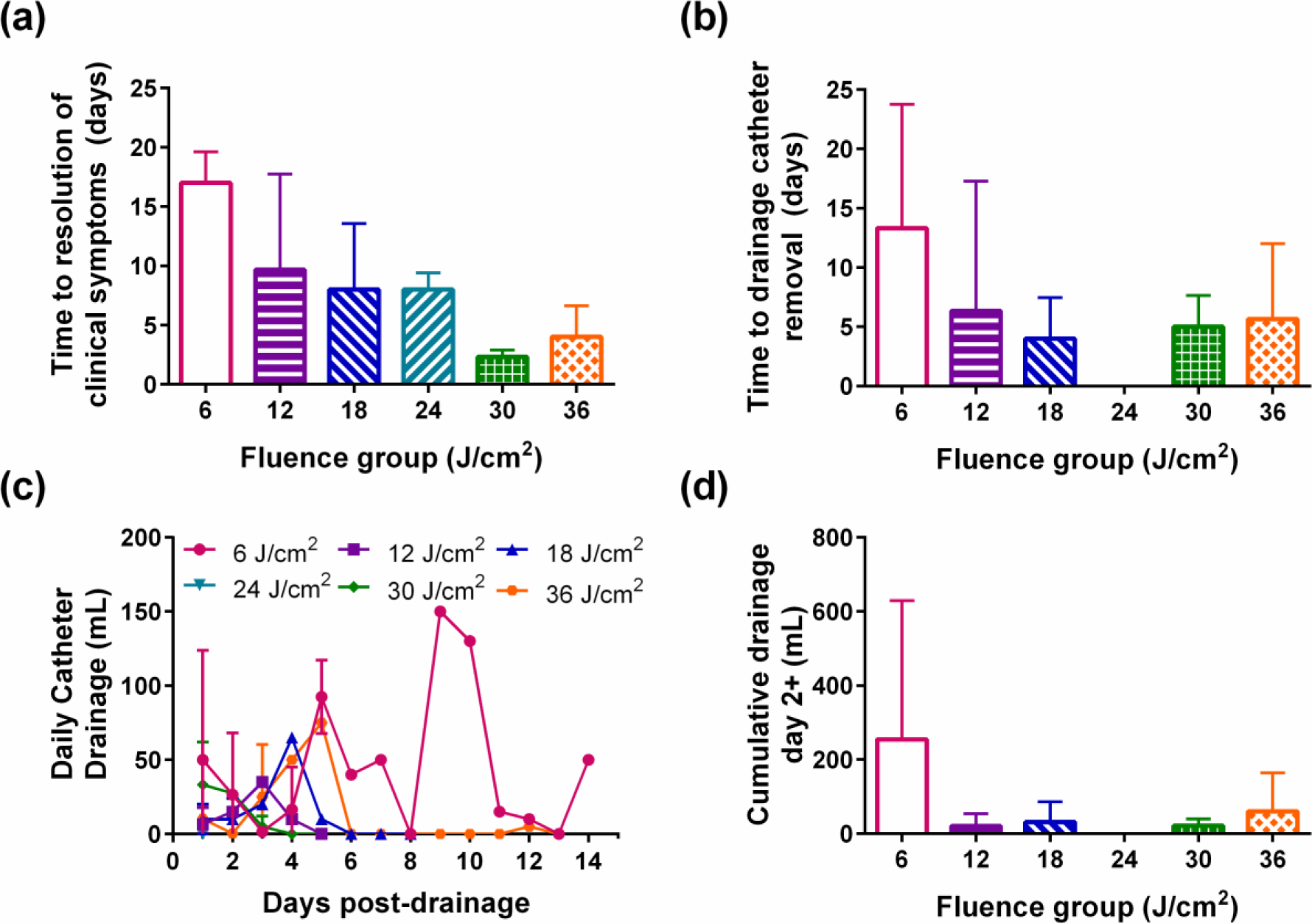
Clinical outcomes separated by fluence group, with values representing mean across subjects within each group and error bars corresponding to standard deviation. (a) Time from drainage/PDT to resolution of clinical symptoms. (b) Time to drainage catheter removal. (c) Daily catheter drainage volume. (d) Cumulative catheter drainage for time points greater than one day after drainage/PDT.

As shown in Figure 3a, time to resolution of clinical symptoms was negatively correlated with fluence (ρ=-0.66, p=0.004). This group difference was significant (p=0.028), with 30 and 36 J/cm^2^ groups showing significant pairwise improvement in symptom resolution compared to the 6 J/cm^2^ group. Controlling for abscess volume, this relationship between fluence and symptom resolution persisted (p=0.002) while abscess volume was not a significant predictor (p=0.085), suggesting that fluence was the main determinant of symptom resolution. This provides evidence for a fluence-dependent response, indicating that PDT may be providing a clinical benefit beyond drainage alone.

As shown in Figure 3b, time to drainage catheter removal post-PDT similarly showed reduction with increasing fluence (ρ=-0.18), although this group difference was not significant (p=0.37). Drainage catheter output decreased after drainage and PDT (Figure 3c). However, there was substantial variability in output on the day following drainage/PDT. We therefore examined cumulative drainage from day two onwards across groups (Figure 3d). There is an apparent reduction in drainage volume with increasing fluence (ρ=-0.18), indicating potential response related to PDT dose. However, this group difference was not significant (p=0.43). When performing linear regression controlling for abscess volume, fluence was still not a significant predictor of time to drain removal (p=0.36) or cumulative drainage volume (p=0.38). These preliminary analyses utilized small sample sizes per study group. It is therefore difficult to conclude whether response is similar between fluence groups, or whether the sample size is insufficient. Future studies will incorporate larger samples sizes to investigate these questions explicitly.

### Microbial Species Isolated

For all abscesses treated, aspirated fluid was retained for identification and antibiotic susceptibility testing. Identified species are tabulated in Table 2. The most common species was *Escherichia coli* (n=5/18). Five abscesses contained antibiotic-resistant bacteria, with four of these containing multi-drug resistant organisms. To determine whether antibiotic-resistant bacteria impacted clinical outcomes, we performed linear regression including measures described in the previous section, with abscess volume and antibiotic resistance as covariates. Antibiotic resistance was not a significant predictor of clinical symptom duration (p=0.37), cumulative drainage volume (p=0.91), or time to drainage catheter removal (p=0.41). Dependence of clinical symptom resolution on fluence was preserved (p=0.0003), indicating that antibiotic resistance did not impact apparent PDT dose response.

**Table 2.**
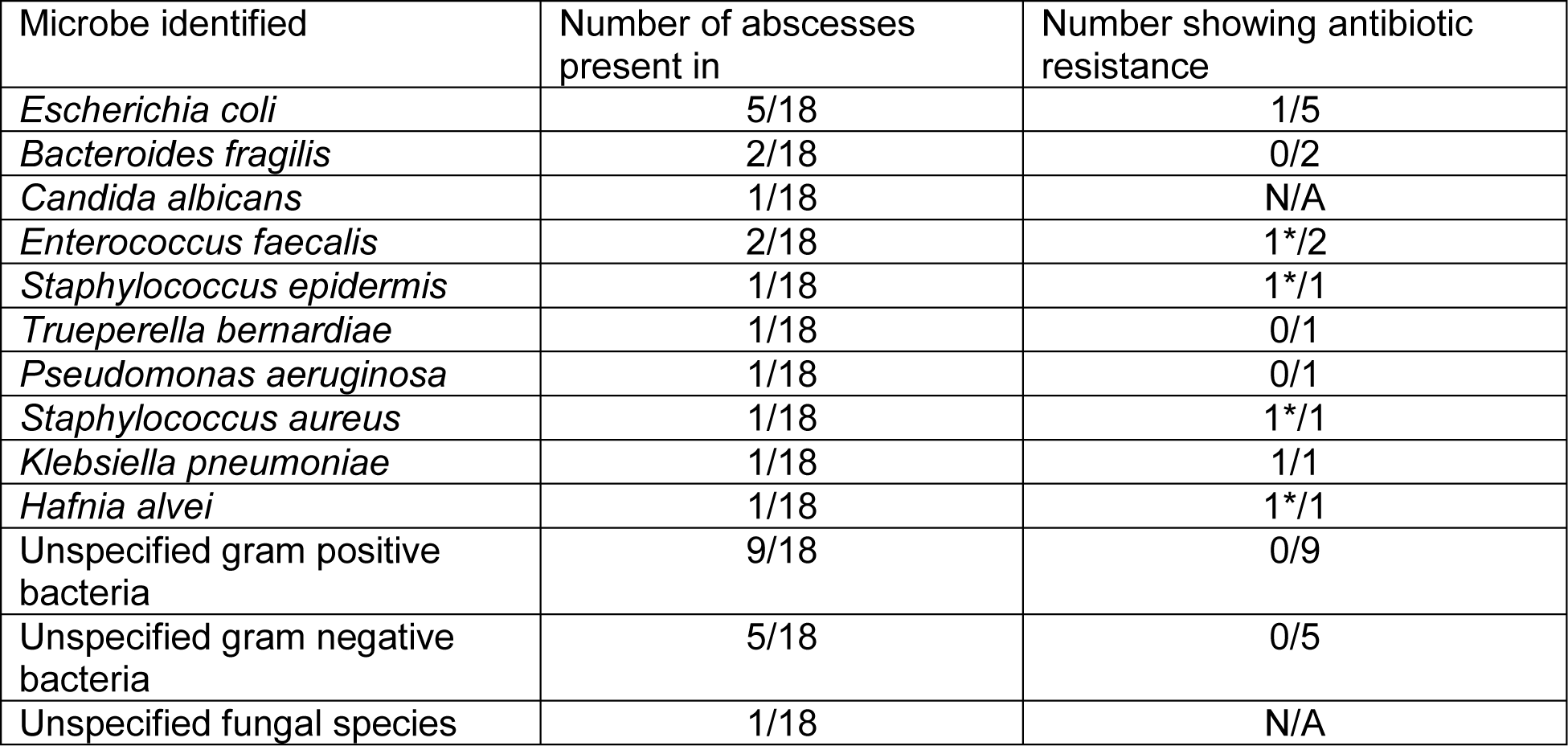
Identified microbes and the number of abscesses in which each species was found, including the number of abscesses in which antibiotic resistant strains were identified. * Multi-drug resistance.

## Discussion

We have demonstrated the safety and feasibility of MB-PDT at the time of image-guided percutaneous abscess drainage. No study-related adverse events were observed, and the procedure was well-tolerated. Based on more rapid resolution of symptoms as a function of fluence, there is preliminary evidence that MB-PDT is effective in eradicating residual infection present in the abscess following drainage. This efficacy will be rigorously examined in future Phase 2 clinical trials.

The expected clinical benefit of MB-PDT is a result of reduction or elimination of bacteria remaining in the cavity following drainage, whether as undrained fluid or bacterial biofilm on the abscess wall. We envision that the eventual use case for MB-PDT would be elimination of drainage catheter placement, with MB-PDT and drainage performed as a single procedure without the need for drainage catheter placement. This would remove catheter monitoring and exchange, reducing discomfort, number of procedures, and healthcare expenses. Future clinical trials will investigate MB-PDT efficacy in detail, before recommending this become routine practice.

MB-PDT could be particularly useful for abscesses containing antibiotic-resistant bacteria. Multiple groups have found antibiotic-resistant bacteria in abscesses^5–7^, and the incidence of these is expected to increase^8^. Even for bacteria that are responsive to antibiotics, drug penetration can prove challenging in the necrotic abscess^25^. Fortunately, MB-PDT is efficacious against antibiotic-resistant bacteria^26–28^, including members of the “ESKAPE” class of pathogens^29^ cultured from human abscess aspirates^6^. In addition to modifying the treatment paradigm for drainage catheter placement, MB-PDT could become a crucial weapon in the fight against antibiotic-resistant bacteria.

Standard of care treatment for abscesses is image-guided percutaneous drainage. While the translation of this to routine practice has significantly reduced morbidity and mortality^3,4^, a number of problems persist. The main issue is variable response rate. Reported clinical success rates range from 77-90%^30,31^. For complicated collections, cure rates as low as 30% have been reported^4^, with deep intramuscular abscesses displaying a 66% response rate^7^. Even for technically successful procedures, catheters can require frequent exchange and extended placement ^30,31^. Additionally, there is lack of consensus on management following drainage^25^.

For these reasons, other adjuvant treatments have been investigated. The most widely investigated has been post-procedure flushing with fibrinolytic agents such as urokinase, which decreases viscosity of purulent fluid present in the abscess^32^. Clinical studies have shown apparent benefit using urokinase flushing^33,34^, though a randomized clinical trial did not show significant differences between urokinase and saline flushing^35^. Fibrinolytic agents have been associated with adverse reactions including hypotension, hypersensitivity reaction, apnea, and bleeding^36,37^. Urokinase treatment also requires flushing multiple times daily for days to weeks. In contrast, MB-PDT is performed at a single time point, and no adverse reactions to MB were observed in the present study.

One of the main criteria excluding potential subjects from enrollment was use of medications with potential MB interactions (Figure 1). Based upon reports that interactions between MB and certain serotonergic psychiatric medications can be related to serotonin syndrome^38^, the Food and Drug Administration issued a drug safety communication related to this interaction^22^. As this was the first study in humans investigating MB-PDT for deep tissue abscesses, we conservatively excluded potential subjects taking these medications. However, these safety concerns are based upon reports where high concentrations (1-8 mg/kg) of MB were delivered intravenously. In contrast, we delivered lower concentrations (0.02-1.5 mg/kg) directly to the cavity, followed by aspiration. 98.7% of MB was recovered in all cases, with 83% of subjects showing complete MB aspiration. The largest concentration of unrecovered MB was 0.1 mg/kg, assuming that unrecovered MB returned to systemic circulation. This concentration was well below that at which drug interactions were reported. For these reasons, exclusion related to serotonergic psychiatric medications will be eliminated in future clinical trials, greatly expanding the eligible patient population.

We acknowledge a number of limitations in the present study. This was an open-label Phase 1 study with limited sample size, so results may be biased by subject characteristics. Further, we excluded potential subjects with abscess diameters >8 cm or multiple abscesses. It is possible that subjects with larger or multiple abscesses may have responded differently. In future Phase 2 studies, we will use a laser with higher maximum output power to allow for treatment of larger abscesses. Additionally, while we tracked surrogates for response to MB-PDT, direct efficacy data were not collected and the study was not adequately powered to determine efficacy. Future Phase 2 studies will be designed to directly evaluate efficacy.

In summary, we have demonstrated that MB-PDT at the time of image-guided percutaneous abscess drainage is safe and feasible. Further, preliminary evidence indicates that MB-PDT may improve the rate of symptom resolution and reduce the need for prolonged drainage catheter placement. We are therefore planning to initiate a Phase 2 clinical trial assessing the efficacy of adding MB-PDT at the time of drainage, compared to drainage alone.

## Data Availability

All data produced in the present study are available upon reasonable request to the authors

## Acknowledgments

The authors would like to thank Dr. Christine Hay for acting as the Independent Safety Monitor, and Dr. Joan Adamo and JoAnne McNamara for regulatory support. This study was supported by grant EB029921 from the National Institutes of Health, and the Harry W. Fischer Research Fund from the Department of Imaging Sciences at the University of Rochester Medical Center.

## Notes

### Competing Interest Statement

The authors have declared no competing interest.

### Clinical Trial

NCT02240498

### Author Declarations

The Research Subjects Review Board of the University of Rochester gave ethical approval for this work.

## References

1 Men, S., Akhan, O. & Köroğlu, M. Percutaneous drainage of abdominal abscess. Eur J Radiol 43, 204–218 (2002).

2 Jaffe, T. A. & Nelson, R. C. Image-guided percutaneous drainage: A review. Abdom Radiol 41, 629–636 (2016).

3 Haaga, J. R. & Nakamoto, D. Computed tomography-guided drainage of intra-abdominal infections. Curr Infect Dis Rep 6, 105–114 (2004).

4 vanSonnenberg, E., Wittich, G. R., Goodacre, B. W., Casola, G. & D’Agostino, H. B. Percutaneous abscess drainage: Update. World J Surg 25, 362–372 (2001).

5 Mezhir, J. J. et al. Current management of pyogenic liver abscess: Surgery is now second-line treatment. J Am Coll Surg 210, 975–983 (2010).

6 Haidaris, C. G. et al. Effective photodynamic therapy against microbial populations in human deep tissue abscess aspirates. Lasers Surg Med 45, 509–516 (2013).

7 Cronin, C. G. et al. Treatment of Deep Intramuscular and Musculoskeletal Abscess: Experience With 99 CT-Guided Percutaneous Catheter Drainage Procedures. American Journal of Roentgenology 196, 1182–1188, doi:10.2214/AJR.09.4082 (2011).

8 Ventola, C. L. The antibiotic resistance crisis: Part 1: Causes and threats. P T 40, 277–283 (2015).

9 Hamblin, M. R. & Hasan, T. Photodynamic therapy: A new antimicrobial approach to infectious disease? Photochem Photobiol Sci 3, 436–450 (2004).

10 Garcez, A. S., Nuñez, S. C., Hamblin, M. R., Suzuki, H. & Ribeiro, M. S. Photodynamic therapy associated with conventional endodontic treatment in patients with antibiotic-resistant microflora: A preliminary report. J Endod 36, 1463–1466 (2010).

11 Liu, Y., Qin, R., Zaat, S. A. J., Breukink, E. & Heger, M. Antibacterial photodynamic therapy: overview of a promising approach to fight antibiotic-resistant bacterial infections. J Clin Transl Res 1, 140–167 (2015).

12 Miranda, R. G. d. & Colombo, A. P. V. Clinical and microbiological effectiveness of photodynamic therapy on primary endodontic infections: A 6-month randomized clinical trial. Clin Oral Investig 22, 1751–1761 (2018).

13 Soukos, N. et al. Photodynamic therapy for endodontic disinfection. J Endod 32, 979–984 (2006).

14 Alvarenga, L. H. et al. Parameters for antimicrobial photodynamic therapy on periodontal pocket—Randomized clinical trial. Photodiagnosis and Photodynamic Therapy 27, 132–136, 10.1016/j.pdpdt.2019.05.035 (2019).

15 Shen, X. et al. Treatment of infected wounds with methylene blue photodynamic therapy: An effective and safe treatment method. Photodiagnosis and Photodynamic Therapy 32, 102051, 10.1016/j.pdpdt.2020.102051 (2020).

16 Cecatto, R. B. et al. Methylene blue mediated antimicrobial photodynamic therapy in clinical human studies: The state of the art. Photodiagnosis and Photodynamic Therapy 31, 101828, 10.1016/j.pdpdt.2020.101828 (2020).

17 Chan, H., Pavelka, M. S. & Baran, T. M. Methylene blue photodynamic therapy of bacterial species found in human abscesses: planktonic, biofilm, and 3D silicone models. in Proc.SPIE 12358. Photonic Diagnosis, Monitoring, Prevention, and Treatment of Infections and Inflammatory Diseases 2023, 1235805, doi:10.1117/12.2648350 (2023).

18 Baran, T. M., Choi, H. W., Flakus, M. J. & Sharma, A. K. Photodynamic therapy of deep tissue abscess cavities: Retrospective image-based feasibility study using Monte Carlo simulation. Med Phys 46, 3259–3267 (2019).

19 Li, Z., Nguyen, L., Bass, D. A. & Baran, T. M. Effects of patient-specific treatment planning on eligibility for photodynamic therapy of deep tissue abscess cavities: retrospective Monte Carlo simulation study. Journal of Biomedical Optics 27, 083007, doi:10.1117/1.JBO.27.8.083007 (2022).

20 Baran, T. M. & Sharma, A. K. Photodynamic Therapy of an Abdominal Abscess at the Time of Percutaneous Drainage. CardioVascular and Interventional Radiology 46, 1292–1294, doi:10.1007/s00270-023-03504-z (2023).

21 Storer, B. E. Design and analysis of phase I clinical trials. Biometrics 45, 925–937 (1989).

22 FDA Drug Safety Communication: Updated information about the drug interaction between methylene blue (methylthioninium chloride) and serotonergic psychiatric medications, <https://www.fda.gov/drugs/drug-safety-and-availability/fda-drug-safety-communication-updated-information-about-drug-interaction-between-methylene-blue> (2011).

23 Hannan, M. N., Sharma, A. K. & Baran, T. M. Preliminary measurements of optical properties in human abscess cavities prior to methylene blue photodynamic therapy. in Proc. SPIE 12359. Optical Methods for Tumor Treatment and Detection: Mechanisms and Techniques in Photodynamic Therapy XXXI, 123590A, doi:10.1117/12.2648453 (2023).

24 Hannan, M. N., Sharma, A. K. & Baran, T. M. First in human measurements of abscess cavity optical properties and methylene blue uptake prior to photodynamic therapy by in vivo diffuse reflectance spectroscopy. Under Review (2023).

25 Wagner, C., Sauermann, R. & Joukhadar, C. Principles of Antibiotic Penetration into Abscess Fluid. Pharmacology 78, 1–10, doi:10.1159/000094668 (2006).

26 Biel, M. A., Pedigo, L., Gibbs, A. & Loebel, N. Photodynamic therapy of antibiotic-resistant biofilms in a maxillary sinus model. Int Forum Allergy Rhinol 3, 468–473 (2013).

27 Garcez, A. S. et al. Effects of antimicrobial photodynamic therapy on antibiotic-resistant Escherichia coli. Photodiagnosis and Photodynamic Therapy 32, 102029, 10.1016/j.pdpdt.2020.102029 (2020).

28 Gulías, Ò., McKenzie, G., Bayó, M., Agut, M. & Nonell, S. Effective Photodynamic Inactivation of 26 Escherichia coli Strains with Different Antibiotic Susceptibility Profiles: A Planktonic and Biofilm Study. Antibiotics 9 (2020).

29 Rice, L. B. Federal funding for the study of antimicrobial resistance in nosocomial pathogens: No ESKAPE. J Infect Dis 197, 1079–1081 (2008).

30 Gee, M. S., Kim, J. Y., Gervais, D. A., Hahn, P. F. & Mueller, P. R. Management of abdominal and pelvic abscesses that persist despite satisfactory percutaneous drainage catheter placement. AJR Am J Roentgenol 194, 815–820 (2010).

31 Marin, D. et al. Percutaneous abscess drainage in patients with perforated acute appendicitis: effectiveness, safety, and prediction of outcome. AJR. American journal of roentgenology 194, 422–429, doi:10.2214/ajr.09.3098 (2010).

32 Park, J. K., Kraus, F. C. & Haaga, J. R. Fluid flow during percutaneous drainage procedures: an in vitro study of the effects of fluid viscosity, catheter size, and adjunctive urokinase. American Journal of Roentgenology 160, 165–169, doi:10.2214/ajr.160.1.8416618 (1993).

33 Haaga, J. R. et al. Intracavitary Urokinase for Enhancement of Percutaneous Abscess Drainage. American Journal of Roentgenology 174, 1681–1685, doi:10.2214/ajr.174.6.1741681 (2000).

34 Lahorra, J. M., Haaga, J. R., Stellato, T., Flanigan, T. & Graham, R. Safety of intracavitary urokinase with percutaneous abscess drainage. American Journal of Roentgenology 160, 171–174, doi:10.2214/ajr.160.1.8416619 (1993).

35 Laborda, A. et al. Percutaneous treatment of intrabdominal abscess: urokinase versus saline serum in 100 cases using two surgical scoring systems in a randomized trial. European Radiology 19, 1772–1779, doi:10.1007/s00330-009-1311-z (2009).

36 Tisdale, J. E., Stringer, K. A., Antalek, M. & Matthews, G. E. Streptokinase-Induced Anaphylaxis. DICP 23, 984–987, doi:10.1177/106002808902301206 (1989).

37 Montserrat, I. et al. Adverse reaction to streptokinase with multiple systemic manifestations. Pharmacy World and Science 17, 168–171, doi:10.1007/BF01879712 (1995).

38 Ng, B. K. W. & Cameron, A. J. D. The Role of Methylene Blue in Serotonin Syndrome: A Systematic Review. Psychosomatics 51, 194–200, 10.1016/S0033-3182(10)70685-X (2010).

